# Plasma proteomics enhances genetic risk prediction and biological understanding of venous thromboembolism

**DOI:** 10.1101/2025.07.09.25330850

**Authors:** Yingyi Mei, Oliver Bundgaard Vad, Astrid Filt Beyer, Jonas Ghouse, Pia Rengtved Lundegaard

## Abstract

Venous thromboembolism (VTE) involves complex genetic and molecular interactions not fully captured by current prediction tools. This study integrated a polygenic risk score (PRS) with plasma proteomics data from 44,138 UK Biobank participants to explore the molecular mechanisms underlying VTE. Associations between PRS_VTE_ and 2,911 plasma proteins were analyzed, identifying 265 significant proteins linked to extracellular matrix organization and transmembrane signaling activity. Cox regressions further identified 354 proteins that significantly associated with incident VTE. Mendelian randomization supported causal relationships for 13 proteins, indicating their potential as therapeutic targets. To improve clinical risk prediction, we developed a protein-based risk score (ProteinRS) using LASSO regression. Here we show that the ProteinRS significantly improves VTE discrimination beyond traditional clinical factors and revealed a three-fold gradient in cumulative VTE risk (Hazard Ratio 0.52 vs 1.49). Our findings demonstrate that integrating plasma proteomics with genetic risk scores provides valuable biological insights and improves prediction of VTE.

## Introduction

Venous thromboembolism (VTE), which includes deep vein thrombosis (DVT) and pulmonary embolism (PE), is a leading cause of morbidity and the third most common vascular cause of death globally, following heart attack and stroke^1^. VTE affects an estimated 350,000 to 600,000 Americans annually, causing over 100,000 deaths, while in the European Union (EU), the yearly disease burden exceeds one million events^2,3^. Although various risk factors and predictors have been identified, and effective preventive therapies are available, the incidence of VTE remains high^4^. This ongoing burden is likely influenced by demographic changes, including an aging population and increasing prevalence of comorbidities^5^.

Despite decades of research, identifying reliable biomarkers and developing effective risk stratification tools for VTE remains a challenge. Previous studies have explored a wide range of candidate biomarkers and predictive models, but many have been limited by small sample sizes, lack of replication, or limited clinical applicability^6–9^. While certain inflammatory or coagulation-related proteins have been shown to associate with VTE risk^10^, their predictive power in clinical settings have often been unsatisfactory^11^. Similarly, clinical risk assessment models, although useful in specific populations, generally lack sensitivity or fail to incorporate biological markers that could reflect individual susceptibility^12^. These limitations emphasize the need for improved biomarkers and innovative approaches to risk stratification that are both biologically informative and clinically viable.

VTE risk is influenced by both acquired and genetic factors, with genetic factors accounting for up to 50-60% of the variance in VTE incidence^13^. Genome-wide association studies (GWAS) have identified multiple common risk variants such as those in F5, F2, and ABO, and enabled the development of polygenic risk scores (PRS) to quantify genetic predisposition^14–16^. However, while PRS can identify individuals with elevated genetic risk, it does not fully reflect the complex and dynamic molecular processes involved in VTE development^17,18^.

Proteomics offers a complementary layer of insight, reflecting real-time biological activity through protein levels. Recent studies have shown that integrating genetic and proteomic data can reveal causal pathways, identify drug targets, and improve disease risk prediction^19–21^.

Moreover, proteomic profiling has demonstrated a strong potential of being a novel standard of disease risk stratification^22–24^. Thus, integrating genomics and large-scale proteomics data holds the potential to provide a more comprehensive understanding of disease development and enable improved approaches to disease risk prediction and stratification.

In this study, we adopted this integrative approach, combining a PRS based on VTE-related genetic variants with plasma proteomics data from more than 44,000 individuals in the UK Biobank. By analyzing 2,911 plasma proteins and constructing a protein-based risk score, this study aimed to deepen the understanding of the molecular pathways underlying VTE and to explore the potential of proteomics to enhance risk prediction and stratification.

## Results

This study included a total of 44,138 individuals of European ancestry. The cohort was composed of 53.7% females and 46.3% males. Participants had a median age of 59 years, and their mean BMI was 27.4 kg/m². A detailed summary of the baseline characteristics is presented in the **Table 1**.

**Table 1.**
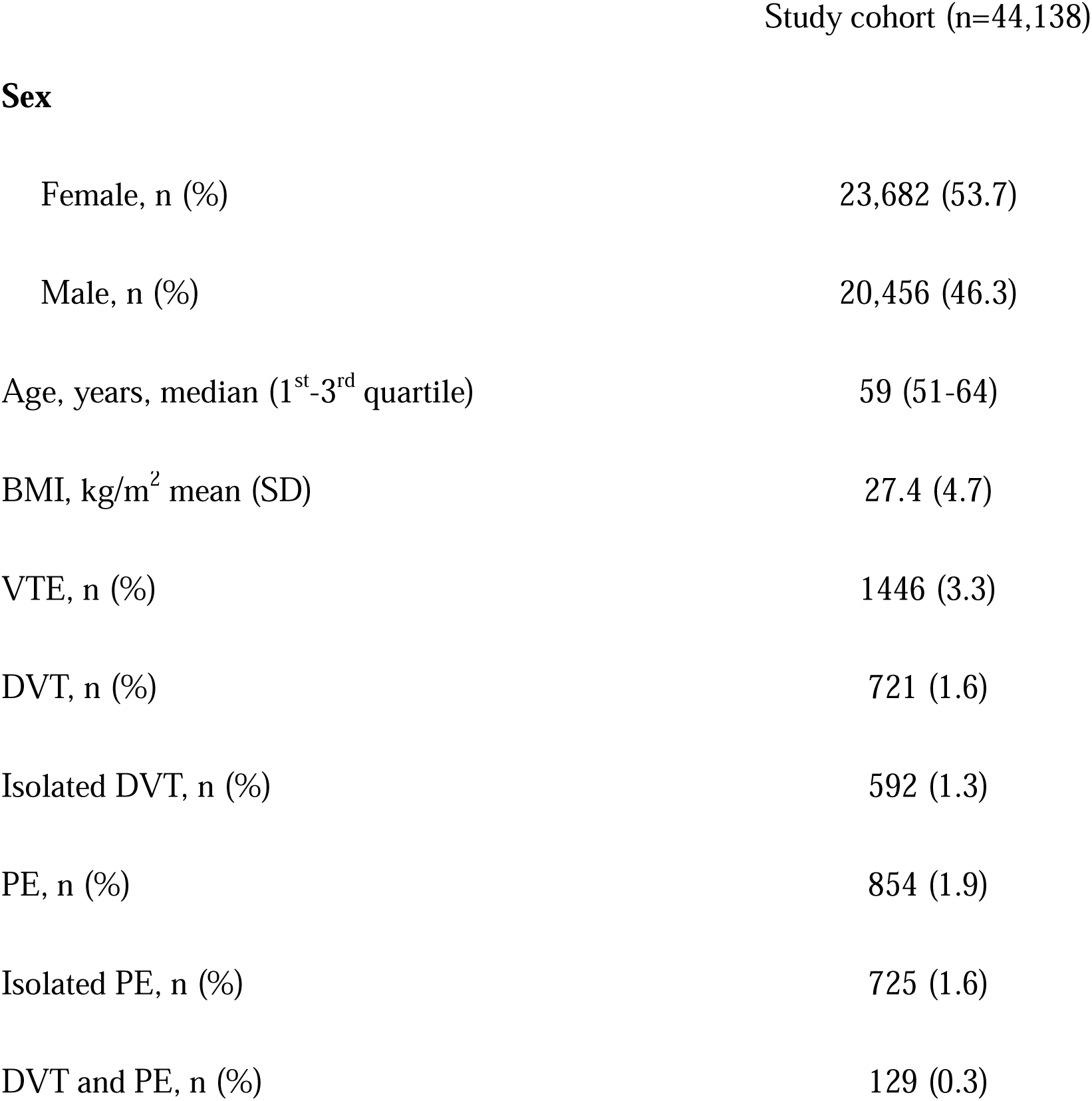
Study Cohort Characteristics. Baseline characteristics of the study cohort (n = 44,138) from the UK Biobank. The median age was 59 years, and 53.7% of participants were female. The mean BMI was 27.4 kg/m² (SD = 4.7). A total of 1,446 participants (3.3%) experienced a venous thromboembolism (VTE) event during follow-up, including isolated deep vein thrombosis (DVT), isolated pulmonary embolism (PE), or both.

### PRS_VTE_ and Protein Expression Level Association

We used multivariable-adjusted linear regression to test the association between PRS_VTE_ and 2911 proteins. We identified 265 proteins with expression levels significantly associated with PRS_VTE_, following FDR correction (*P* < 0.01). The associations are visualized in **Figure 1A**. The most significant proteins were ABO, SELE, and CD209. The proteins with the largest effect estimates were ABO, MUC2 and CLEC4M (up-regulated), and ALPI and SELE (down-regulated). The full list of significant proteins is provided in **Supplementary Table 1**.**Pathway Enrichment Analysis**

**Figure 1:**
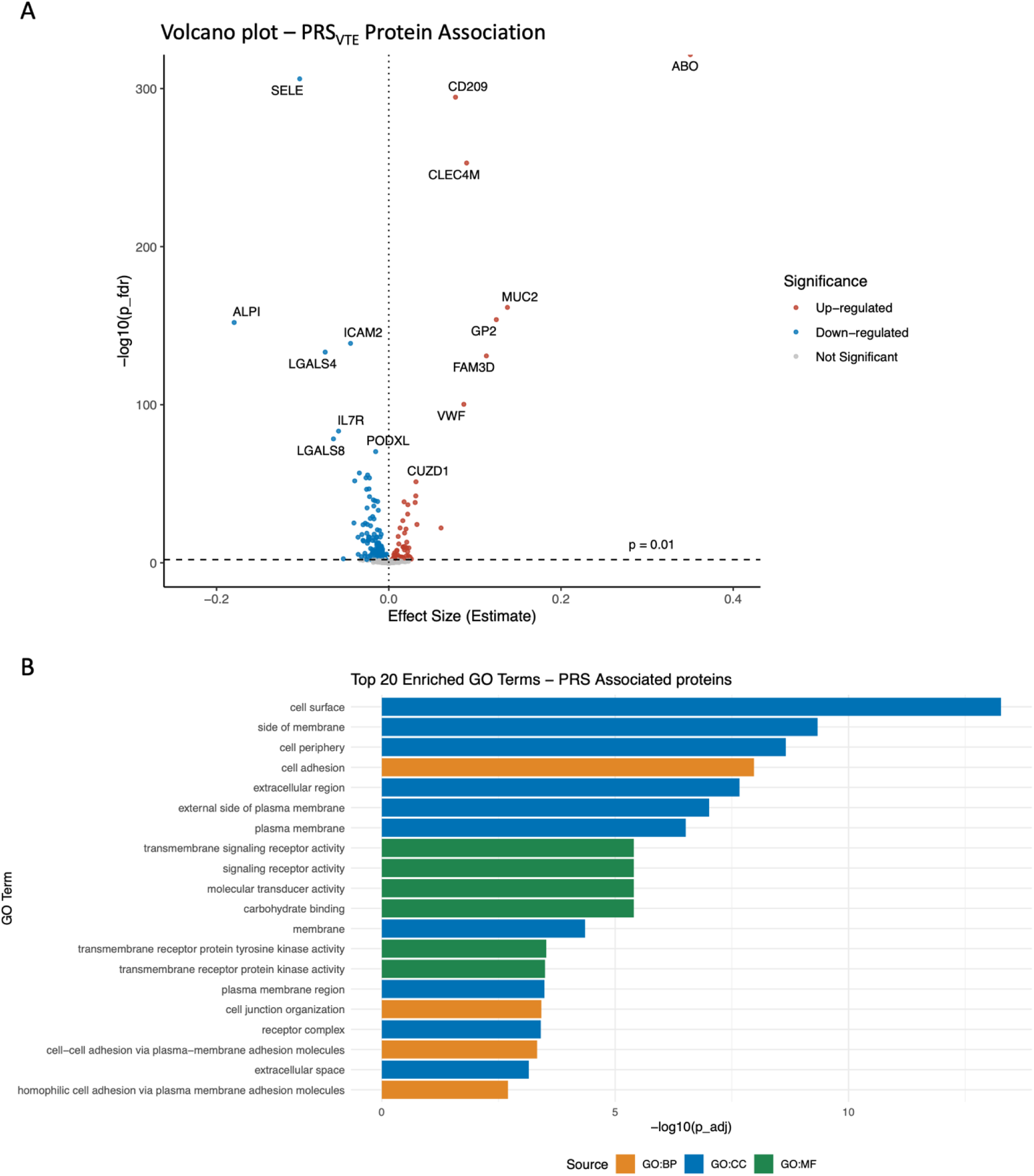
PRS_VTE_-associated proteins: volcano plot and pathway enrichment. **A**: Volcano plot showing significant PRS_VTE_-proteins associations. Red and blue points represent significantly up-and down-regulated proteins respectively (FDR-adjusted p < 0.01). **B:** Top 20 enriched Gene Ontology (GO) terms for PRS_VTE_-associated proteins, categorized as Biological Process (BP), Cellular Component (CC), and Molecular Function (MF).

The pathogenesis of VTE is traditionally explained by Virchow’s triad, which includes venous stasis, endothelia injury, and hypercoagulability^25,26^. Accordingly, we expect to see enrichment pathway related to platelet activation, coagulation cascade, and hemostasis. The Gene Ontology (GO) enrichment analysis of the 265 PRS_VTE_-associated proteins confirmed this. The top 20 enriched GO terms (**Figure 1B**) included pathways linked with cellular locations such as the cell surface, side of membrane, and extracellular region. Notably enriched biological processes involved cell adhesion, cell junction organization, cell-cell adhesion via plasma-membrane adhesion molecules, and homophilic cell adhesion via plasma membrane adhesion molecules. For molecular functions, prominent terms included transmembrane signaling receptor activity, transmembrane receptor protein kinase activity, transmembrane receptor protein tyrosine kinase activity, signaling receptor activity, molecular transducer activity, and carbohydrate binding.

Detailed results are summarized in **Supplementary Table 2.** Tissue-expression enrichment showed that the these proteins are most expressed in omentum, gut epithelium (colon, ileum, small intestine, stomach) and liver, tissues that secrete many circulating coagulation and adhesion proteins (adjusted p < 1 × 10 ¹ ; **Supplementary Table 3**).

### Proteins Associated with Incident Disease

The median follow-up time was 13.8 years (1^st^-3^rd^ quartile: 13.0-14.5 years). Over the follow-up period, 1446 (3.3%) individuals developed VTE. Within these groups, 725 (1.6%) were identified as having isolated PE, 592 individuals (1.3%) were classified as having isolated DVT, and 129 (0.3%) experienced PE with a concurrent DVT diagnosis. A total of 4,850 participants (11%) died during the follow-up period.

We tested the associations of 2911 proteins with the incident VTE by multivariable-adjusted Cox proportional hazards regression model. **Figure 2** shows the correlation between log hazard ratios for incident VTE and effect estimates from PRS_VTE_-protein regression. There were 354 proteins significantly associated with the incidence of VTE risk after FDR correction. **Supplementary Table 4** contains the full list of proteins with significant association with incident VTE. Fifty-six proteins (**Supplementary Table 5**) showed significant associations with both incident VTE risk and PRS_VTE_, including ABO, MUC2, GP2, CLEC4M, CD209, ADAMTS13, and ALPI. Among them, 49 exhibited consistent directional trends between HR values and expression estimates, showing a concordance between genetically predicted changes in protein expression and their associated risk for VTE. Other proteins, including FAM3D, SELE, and LGALS4, were significantly associated exclusively with PRS_VTE_. Conversely, proteins like EGFR, ITGAV, PLAUR, and SERPINA3 demonstrated significant associations only with incident VTE. In penalized smoothing spline models, we observed three proteins with evidence of significant non-linear relationships with VTE (FDR < 0.01), including CD300LF, EDA2R, and ICAM3 (**Extended Data Fig. 1**). We observed U-shaped relationships for EDA2R, while ICAM3 exhibited increasing HRs at high expression levels.

**Figure 2:**
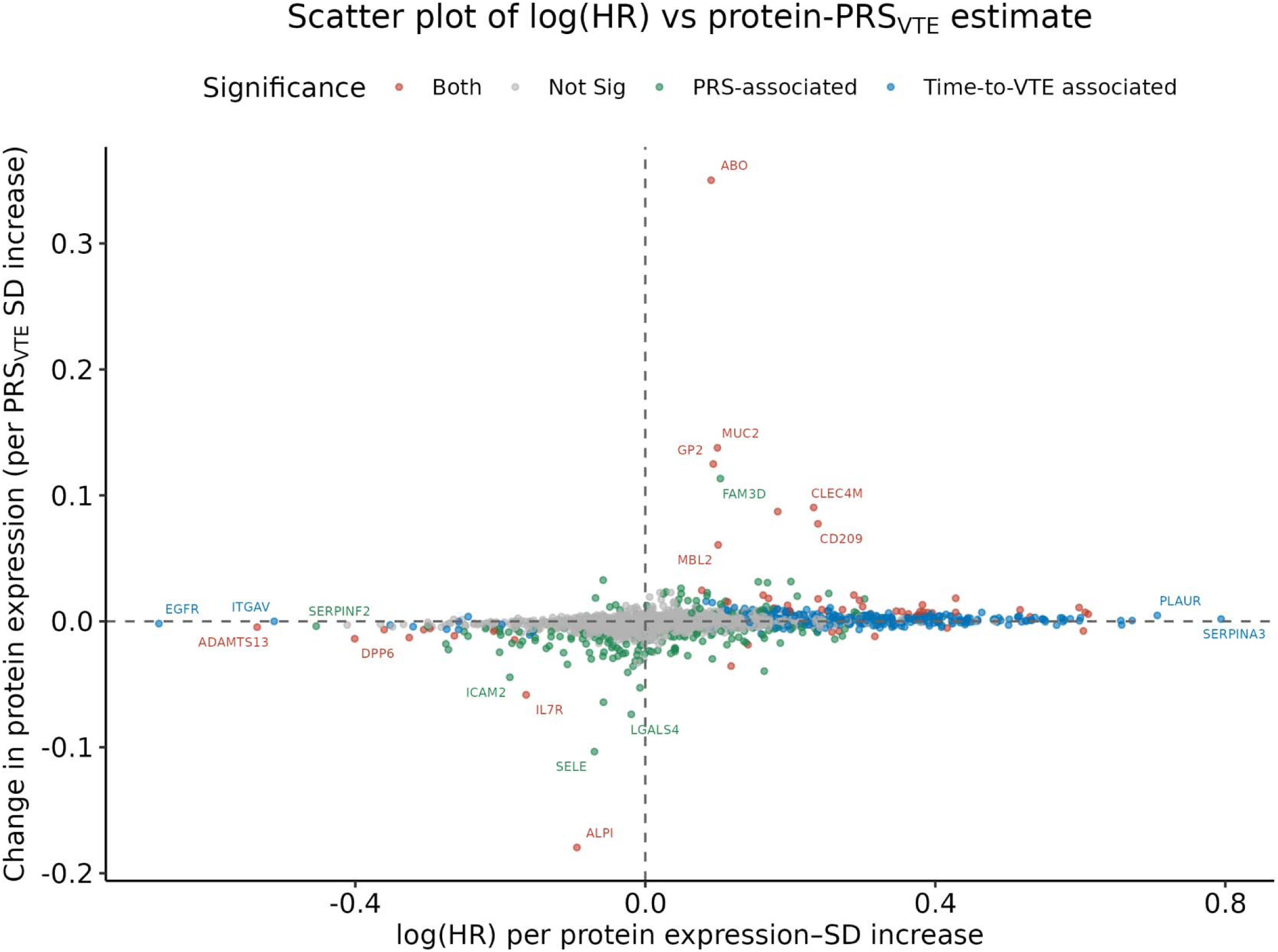
Scatter plot showing the relationship between log hazard ratios (HR) derived from Cox regression analyses and protein expression changes (Estimate) from linear regression analyses. Proteins are colored according to their statistical significance: red indicates proteins significantly associated with both time-to-VTE risk and PRS_VTE_, green indicates significant association exclusively with PRS_VTE_, blue indicates significant association with only time-to-VTE risk, and grey represents proteins without significant associations. The dashed vertical line at log HR = 0 corresponds to HR = 1 (no association with VTE risk), while the dashed horizontal line at 0 indicates no change in protein expression.

### Mendelian Randomization (MR) Analysis

We conducted a bidirectional two-sample MR analysis to investigate potential causal associations between 56 proteins and VTE risk. These 56 proteins were significantly associated with both PRS_VTE_ and incident VTE in our observational analyses shown above. Results of significant associations after correction for multiple comparisons (FDR < 0.05) are showed in **Figure 3** (protein to VTE direction) and **Figure 4** (VTE to protein direction). Comprehensive results for all 56 proteins are detailed in **Supplementary Tables 7 and 8**, as well as **Extended Data Figures 2 and 3**.

**Figure 3.**
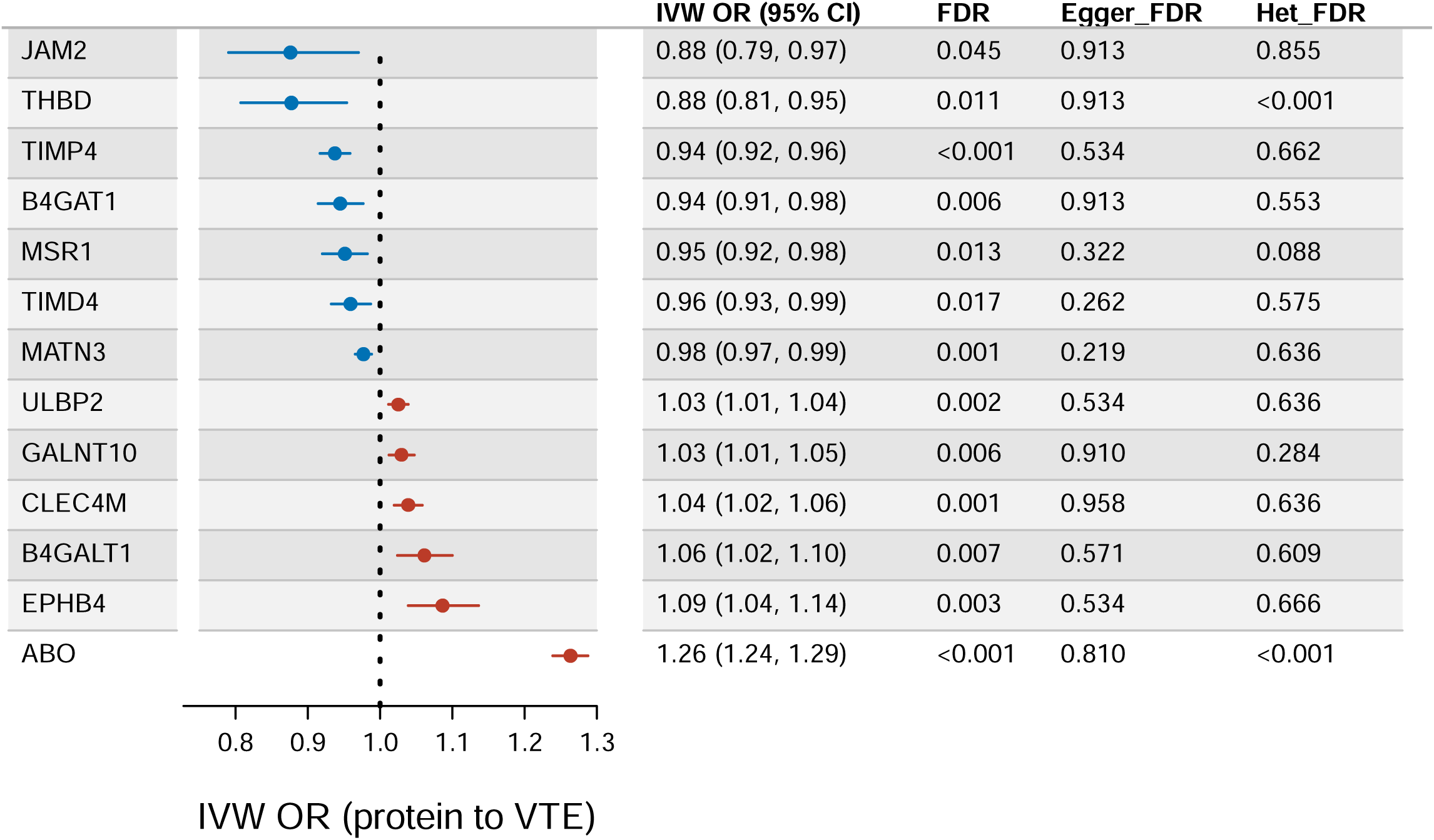
Mendelian randomization results for significant protein associations with VTE risk. Odds ratios (ORs) and 95% confidence intervals (CIs) from inverse-variance weighted (IVW) MR analyses represent causal estimates of protein levels on VTE risk. Blue indicates a higher protein level associated with lower VTE risk, red indicates a higher protein level associated with higher VTE risk. Cochran’s Q *P* (heterogeneity) and Egger intercept *P* (pleiotropy) are shown. *P* values were adjusted for multiple tests using FDR correction.

**Figure 4.**
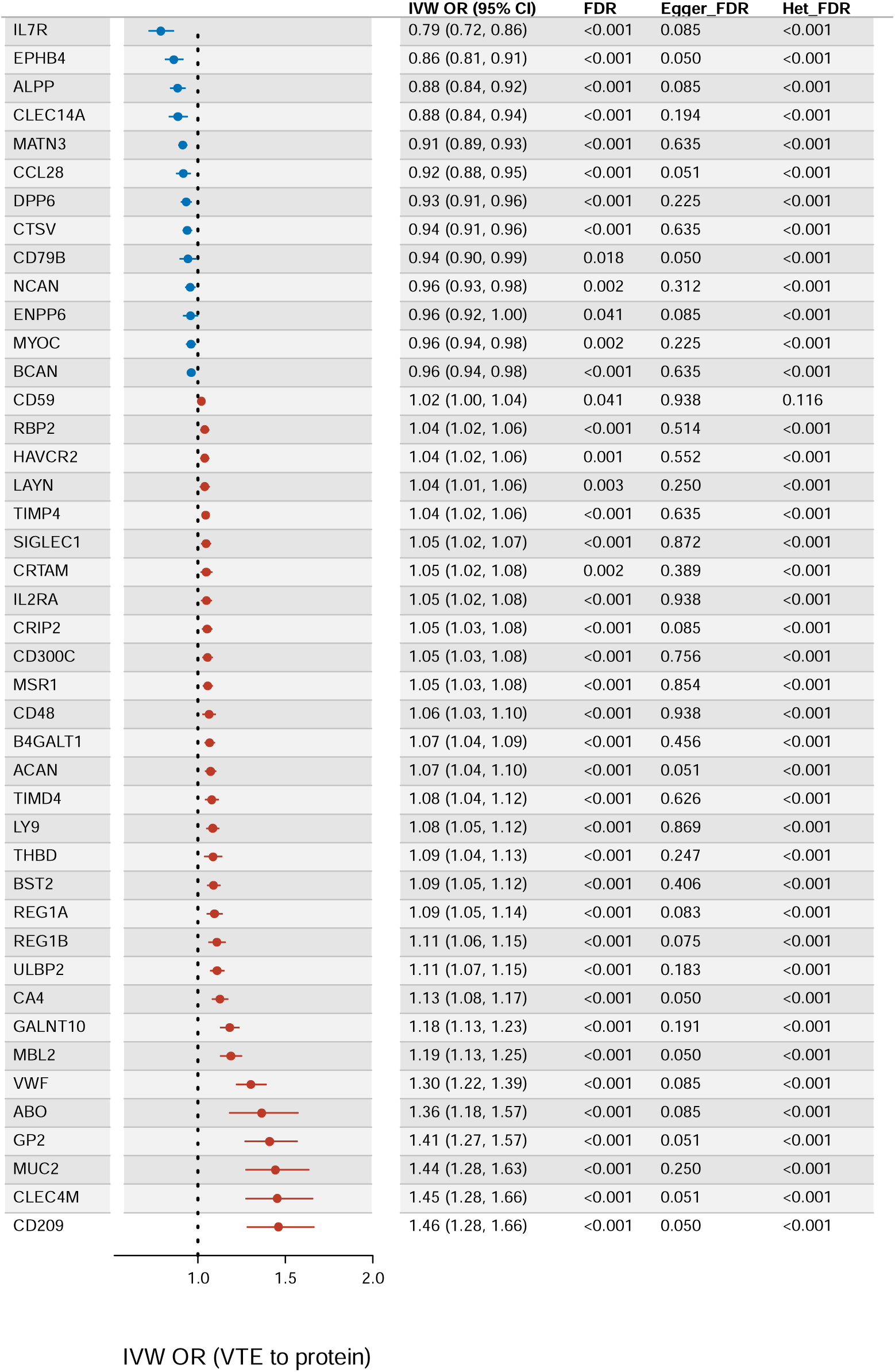
Mendelian randomization results for significant associations of VTE genetic liability with protein levels. Odds ratios (ORs) and 95% confidence intervals (CIs) from inverse-variance weighted (IVW) MR analyses represent causal estimates of VTE genetic predisposition on protein expression levels. Blue dots indicate proteins with significantly lower levels associated with increased VTE genetic liability, red dots indicate proteins with significantly higher levels associated with increased VTE genetic liability. Cochran’s Q *P* (heterogeneity) and Egger intercept *P* (pleiotropy) are shown. *P* values were adjusted for multiple tests using FDR correction.

In the forward direction (protein to VTE), we identified 13 proteins significantly associated with VTE after FDR correction. Specifically, we observed a protective effect of JAM2, THBD, TIMP4, B4GAT1, MSR1, TIMD4, and MATN3, with the inverse-variance weighted (IVW) odds ratio (OR) ranging from 0.88 to 0.98. On the contrary, higher levels of ABO, EPHB4, B4GALT1, CLEC4M, GALNT10, and ULBP2 were associated with an increased VTE risk (IVW OR range: 1.03 to 1.26). Comparison with the observational analyses showed full directional concordance for six proteins: B4GAT1 was consistently protective (MR OR 0.94; Cox HR 0.69), while ULBP2, GALNT10, CLEC4M, B4GALT1, EPHB4, and ABO were consistently risk-enhancing (MR OR > 1; Cox HR >). No evidence of horizontal pleiotropy was noted for all tested proteins. Cochran’s Q indicated evidence of heterogeneity for THBD and ABO only (**Figure 3**).

In the reverse direction (VTE to protein), genetic predisposition to VTE was significantly associated with levels of 42 proteins after FDR correction. The strongest inverse associations were observed with IL7R (OR, 0.79 [95% CI, 0.72-0.86]; FDR < 0.001) and EPHB4 (OR, 0.86 [95% CI, 0.81-0.91]; FDR < 0.001). The strongest positive associations included CD209 (OR, 1.46 [95% CI, 1.28-1.66]; FDR < 0.001) and CLEC4M (OR, 1.45 [95% CI, 1.28-1.66]; FDR < 0.001). MR-Egger intercept indicates no horizontal pleiotropy for most associations, with evidence of potential pleiotropy for EPHB4, CD79B, CA4, and CD209 (Egger FDR ≤ 0.05). However, except for CD59, Cochran’s Q identified heterogeneity for all these significant proteins **(Figure 4).**

### Protein-based Risk Score (ProteinRS)

Using the optimal regularization parameter selected via tenfold cross-validation, we built three LASSO regression models (protein-only, clinical-only, and combined). In the protein-only model, we identified a set of proteins significantly associated with incident VTE risk from the high-dimensional dataset. **Figure 5A** illustrates the protein coefficients from the optimal protein-only LASSO model. There were 60 proteins with non-zero coefficients (**Supplementary Table 9**).

**Figure 5.**
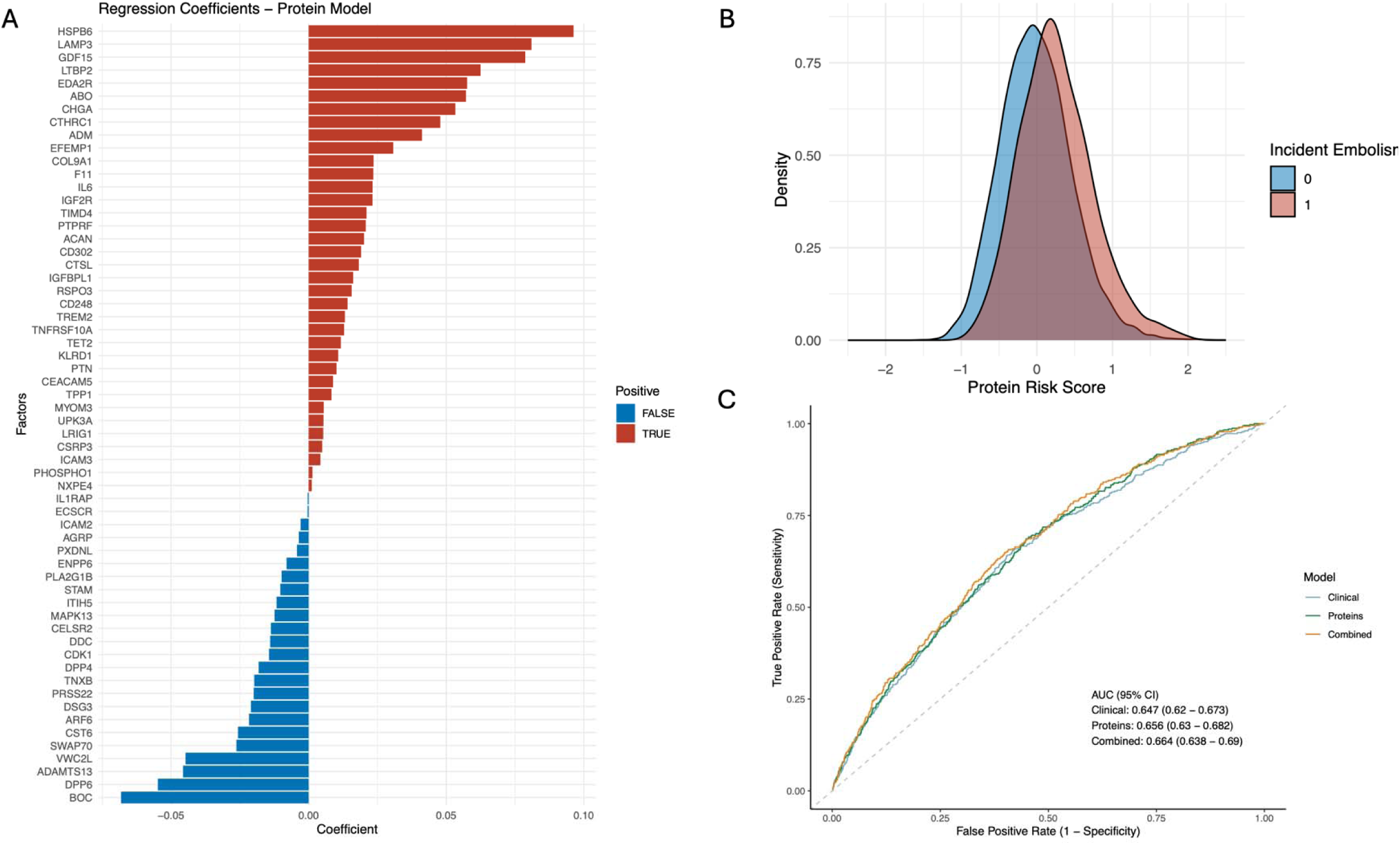
Development and Validation of Protein-based Risk Score (ProteinRS). **A:** LASSO regression coefficients of proteins identified from the optimal protein-only model associated with incident VTE risk. Positive coefficients (red) indicate proteins associated with increased risk, while negative coefficients (blue) indicate protective associations. **B**: Density distribution of ProteinRS in the testing set, categorized by incident VTE status. **C**: ROC curves comparing the predictive performance of clinical-only, protein-only, and combined models in predicting incident VTE. The AUC and corresponding 95% confidence intervals are provided for each model.

Proteins such as HSPB6, LAMP3, GDF15, and LTBP2 were key factors contributing positively to VTE risk, whereas proteins like BOC, DPP6, and ADAMTS13 demonstrated negative associations. We then computed a ProteinRS for each individual in the testing set (**Supplementary Table 10**). The distribution of ProteinRS differed significantly between VTE cases and controls, as visualized by density plots (**Figure 5B**). VTE cases had a higher mean ProteinRS (mean = 0.261, SD = 0.482) than controls (mean = –0.009, SD = 0.470), and this difference was statistically significant (P < 2.2×10□¹□, Wilcoxon rank-sum test)

Receiver operating characteristic (ROC) analyses compared the predictive performance across the models. The protein-only model achieved an area under the curve (AUC) of 0.656 (95% CI: 0.630–0.682), slightly higher than the clinical-only model, which had an AUC of 0.647 (95% CI: 0.620–0.673). The combined model integrating both clinical and protein variables demonstrated better predictive ability, achieving the highest AUC of 0.664 (95% CI: 0.638–0.690) (**Figure 5C**). Pairwise comparisons using DeLong’s test revealed statistically significant improvements in predictive performance for the combined model compared to the clinical-only model (P = 0.02405).

Incidence rates of VTE, calculated per 1,000 person-years, demonstrated a clear positive correlation with ProteinRS percentiles. Participants in higher risk percentiles displayed markedly higher cumulative risk (**Figure 6A**). Risk stratification by percentiles of ProteinRS demonstrated a significant gradient in VTE risk. Individuals with ProteinRS above the 80^th^ percentile had a substantially increased rate of VTE compared to the reference (20-80^th^ percentile) group, with an HR of 1.49 (95% CI: 1.17-1.90, P = 0.00114). Conversely, individuals below 20^th^ percentile showed significantly reduced VTE risk (HR, 0.52, [95% CI, 0.34-0.79], P = 0.00251) (**Figure 6B**).

**Figure 6.**
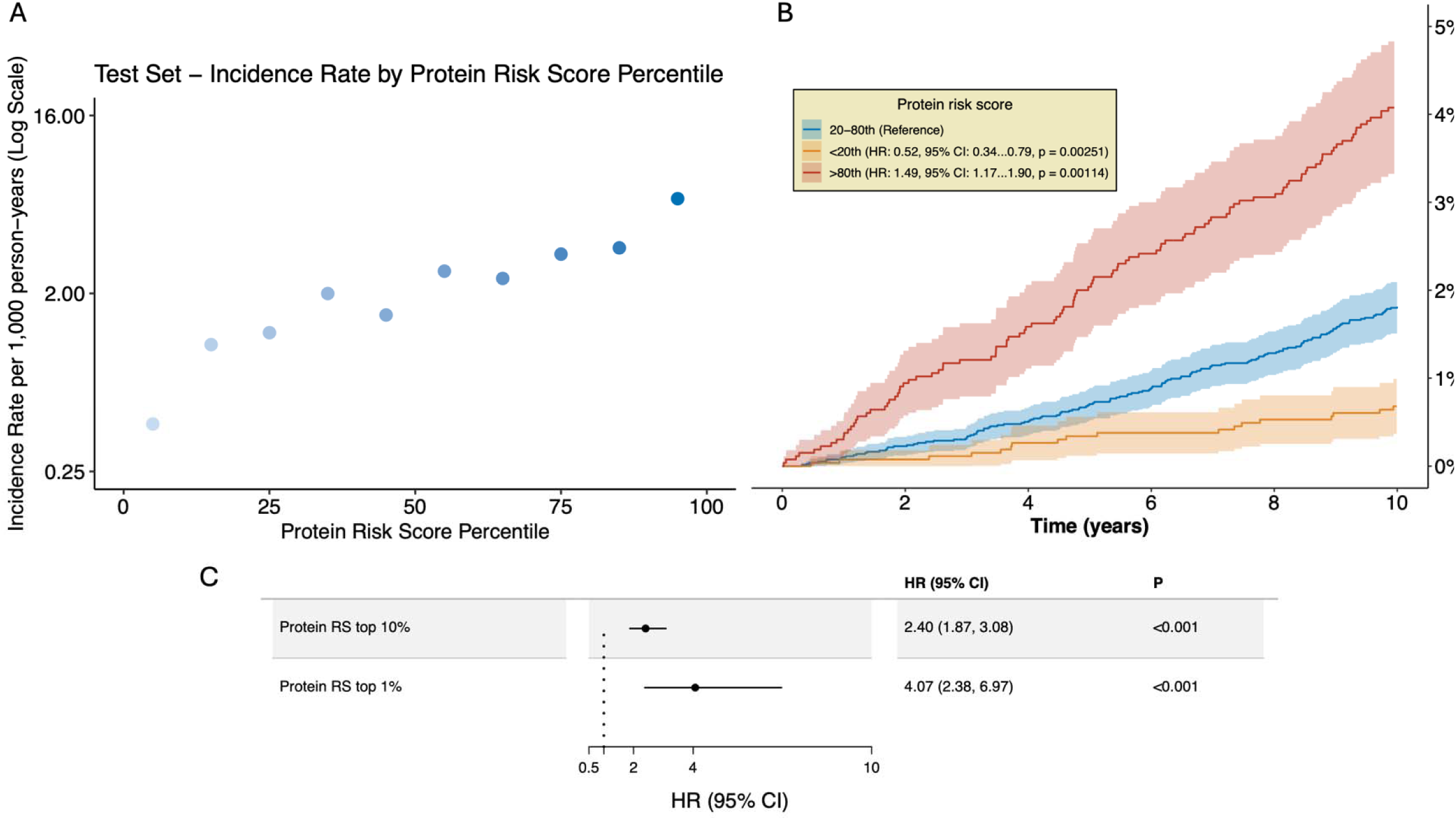
Risk Stratification and Incidence Analysis using ProteinRS. **A:** Incidence rates of VTE per 1,000 person-years plotted by ProteinRS percentiles on a logarithmic scale. **B:** Cumulative incidence curves of VTE events across three percentile-based groups: <20^th^ percentile, 20-80^th^ percentile (reference), and >80^th^ percentile. Cox-regression derived hazard ratios with 95% confidence intervals and corresponding *P* values are reported. **C:** Forest plot displaying hazard ratios for VTE risk among individuals in the top 10% and top 1% ProteinRS groups. Hazard ratios, 95% confidence intervals, and *P* values are presented, demonstrating significantly elevated risk within these high-risk groups.

Compared with the rest of the cohort, individuals in the top 10% of ProteinRS showed a significantly higher hazard of VTE (HR: 2.40, 95% CI: 1.87–3.08, p < 0.001). This association was even more pronounced when comparing individuals in the top 1% of ProteinRS with the remainder of the cohort (HR: 4.07, 95% CI: 2.38–6.97, p < 0.001) (**Figure 6C**).

## Discussion

### Overview of Key Findings

Our integrative analysis of 44,138 UK Biobank participants provides novel insight into the molecular basis of VTE by identifying 265 proteins significantly associated with the PRS_VTE_. Among these proteins, key markers such as ABO, SELE, and CD209 exhibited particularly strong associations, highlighting their potential roles as intermediaries that bridge genetic predisposition and clinical manifestations of VTE. Pathway enrichment analysis further suggested that disruptions in extracellular adhesion and cellular signaling pathways contribute to VTE pathogenesis. Complementary analyses using Cox regression identified 354 proteins predictive of incident VTE events, including 56 overlapping proteins shared with PRS_VTE_ association findings, reinforcing their functional relevance in disease progression. Extending from these two-steps’ observational analysis, the bidirectional two-sample Mendelian randomization further supported causal relationships between the 56 proteins and VTE, identifying 13 proteins whose levels significantly influenced VTE risk. Conversely, genetic predisposition to VTE significantly influenced the expression levels of 42 proteins. Finally, the development of a protein-based risk score derived from 60 significant proteins enhanced VTE prediction beyond traditional clinical models, providing a novel method for improved clinical risk stratification and potentially guiding individualized prevention interventions. Collectively, these findings not only advance our biological understanding of VTE pathogenesis but also hold promise for future clinical translation, particularly in the context of targeted biomarkers discovery and personalized risk stratification strategies.

### Proteins Associated with Both PRS_VTE_ and Incident VTE

Comparing our findings with existing literature reveals insights into the molecular mechanisms underlying VTE, confirming several known pathways and raising novel questions. Among the 56 proteins significantly associated with both PRS_VTE_ and incident VTE, several have previously been implicated in embolism-related disease pathophysiology. ADAMTS13, a metalloprotease responsible for cleaving prothrombotic ultra-large VWF multimers, has been shown to protect against thrombotic events like thrombotic thrombocytopenic purpura (TTP)^27–32^. Our results align with this protective role, showing reduced VTE risk associated with increased ADAMTS13 levels (HR=0.59, 95% CI: 0.48–0.71; P<0.001), together with its downregulation in genetically predisposed individuals. Conversely, VWF which mediates platelet adhesion and aggregation, was positively associated with genetic VTE predisposition and incident VTE (HR=1.20, 95% CI: 1.13–1.28; P<0.001), reinforcing its established thrombotic role^27,28^. TIMD4 was upregulated in those genetically predisposed to VTE, with an HR of 1.42 (95% CI, 1.23–1.65; P<0.001).

TIMD4 is known for its role in macrophage-driven apoptotic cell clearance and influences inflammatory and thrombotic pathways and has been implicated in cardiovascular disease progression^33–37^. Macrophages, which express TIMD4, are increasingly recognized as key players in thrombus resolution and immune modulation^38,39^. This supports a potential role for TIMD4 in inflammation in VTE. Interestingly, our findings on CD59 contrast with previous reports. CD59 is a regulator of the complement system that prevents membrane attack complex formation. It has traditionally been considered protective against thrombosis, as shown by the elevated thrombotic risk observed in CD59-deficient conditions, such as paroxysmal nocturnal hemoglobinuria (PNH)^40–46^. However, in our analysis, it was overexpressed in individuals with high PRS_VTE_ and was associated with increased rates of incident VTE (HR=1.7, 95% CI: 1.36– 2.13; P=0.011). This discrepancy may reflect compensatory mechanisms responding to increased complement activity, endothelial injury, or inflammation rather than CD59 directly promoting thrombosis. These findings both confirm known associations and uncover novel interactions, highlighting the complexity of VTE biology and the value of our systematic proteomic-genetic approach.

### Mendelian Randomization Highlights Potential Causal Proteins

In our subsequent MR analysis of 56 proteins, we identified six proteins (ABO, EPHB4, B4GALT1, CLEC4M, GALNT10, and ULBP2) that were associated with increased VTE risk. These findings were consistent with the observational analyses previously, supporting a causal role for their circulating levels in VTE. EPHB4 (MR OR = 1.09, FDR = 0.003; Cox HR = 1.83, FDR < 0.001), a receptor tyrosine kinase critical for vascular development and venous endothelial differentiation, implicating its role in structural valve dysfunction and thrombosis driven by venous stasis^47–50^. Interestingly, reverse MR analysis for EPHB4 (OR = 0.86, FDR < 0.001) suggested a feedback loop where pro-thrombotic genotypes down-regulate endothelial EPHB4, potentially exacerbating valve dysfunction. Yuan et al. also identified EPHB4 as a novel causal target using a cohort and MR approach, further support EPHB4’s potential role in VTE^51^. CLEC4M (MR OR = 1.04, FDR < 0.001; Cox HR = 1.26, FDR = 0.001) functions by binding and internalizing VWF and factor VIII (FVIII), thus strongly influencing their circulating levels^52^. While VWF and FVIII are two well-characterized risk factors for VTE^53,54^, impaired CLEC4M function may contribute to elevated VTE risk through disrupted clearance of these prothrombotic proteins^55^. Additionally, the glycosyltransferases B4GALT1 (MR OR = 1.06, FDR = 0.007; Cox HR = 1.48, FDR < 0.001) and GALNT10 (IVW OR = 1.03, FDR = 0.006; Cox HR = 1.35, FDR = 0.006) could influence VTE risk by modifying the glycosylation patterns of VWF^56,57^. ULBP2 (IVW OR = 1.03, FDR = 0.002; Cox HR = 1.53, FDR < 0.001), a stress-induced ligand connecting innate immunity and coagulation, potentially indicated the role of immunothrombosis driven by NK-cell-mediated endothelial injury^58–60^.

By contrast, seven other proteins (JAM2, THBD, TIMP4, B4GAT1, MSR1, TIMD4, and MATN3) showed protective MR associations. Among then, only B4GAT1 remains consistently protective across all analyses (MR OR 0.94; Cox HR 0.69). Its vascular expression and role in glycan priming may influence the clearance of pro-thrombotic proteins like VWF and factor VIII, offering a potential mechanism for its anti-thrombotic effect^61–63^. THBD (MR OR = 0.88, FDR = 0.011; Cox HR = 1.68, FDR < 0.001) activates protein C to exert anti-thrombotic effects, and its elevated plasma levels likely reflect reactive endothelial shedding in response to thrombin generation^64–66^. TIMP4 (MR OR = 0.94, FDR < 0.001; Cox HR = 1.22, FDR = 0.007), a platelet inhibitor of metalloproteinases^67^, genetically appears protection but higher baseline levels may indicate platelet activation appear before thrombosis. Macrophage-associated proteins MSR1 (MR OR = 0.95, FDR = 0.012; Cox HR = 1.28, FDR < 0.001) and TIMD4 (MR OR = 0.96, FDR = 0.004; Cox HR = 1.34, FDR < 0.001) are macrophage receptors that help clear apoptotic cells and restrain vascular inflammation^35,68^. The similar discordance of these two proteins may suggesting that individuals with ongoing vascular inflammation have higher plasma levels even though lifelong genetically higher expression is protective. These contrasts illustrated how MR analysis could help separate proteins that cause thrombosis from those that are elevated because early and silent disease processes are already under way.

Bidirectional MR analyses revealed directional complexities and potential feedback mechanisms involving several proteins, notably EPHB4 and B4GAT1. MR-Egger test found no evidence of directional pleiotropy, while Cochran’s Q showed significant heterogeneity for THBD and several reverse MR results, implying that some genetic instruments may affect VTE through pathways unrelated to the target proteins. Thus, proteins exhibiting concordant signals and low heterogeneity, like CLEC4M, have stronger causal inference support.

### Protein-Based Risk Score and Predictive Utility

The final stage of our analysis focused on building three predictive models using LASSO regression: a clinical model, a protein-only model, and a combined model integrating both clinical and proteomic variables. The protein coefficients from the LASSO-selected protein model were then used to construct a ProteinRS for each individual in a test subset. Both the protein and combined model performed better than the clinical model, with the combined model showing a statistically significant improvement. This result suggests that plasma proteomics can improve prediction beyond traditional clinical factors. Notably, the 60 proteins selected by the LASSO model highly overlapped with those significantly associated with either PRS_VTE_ or incident VTE, supporting their potential biological relevance. The ProteinRS showed modest discriminative ability and improved risk separation compared to the clinical model alone. Density plots showed a clear separation between cases and controls, while individuals with higher scores exhibited increased incidence rates and cumulative hazard. Risk stratification by ProteinRS percentiles confirmed its strong association with VTE incidence. Individuals in the highest ProteinRS group (>80^th^ percentile) had approximately 1.5-fold elevated risk, while those in the lowest group (<20^th^ percentile) had nearly 50% reduced risk compared to the intermediate group (20-80^th^ percentile). These findings reinforce the clinical potential of ProteinRS for identifying individuals at both elevated and reduced risk of VTE. Overall, our findings demonstrated that proteomics not only provides insights into disease mechanisms but also substantially improves risk prediction and stratification when integrated with clinical data.

### Limitations

This study has several limitations that should be considered when interpreting the findings. First, we restricted our analysis to participants of European ancestry to ensure consistency with the underlying GWAS used to construct the PRS_VTE_ and to reduce risk of bias from population stratification. As a result, our findings may not be generalizable to individuals of non-European ancestry. Second, although multiple testing corrections were applied, the high-dimensional nature of proteomic data introduces the potential for false-positive associations. Third, the UK Biobank is known to exhibit a “healthy volunteer bias,” as participants tend to be healthier, and better educated than the general population^69^. This selection bias may lead to underestimation of disease prevalence or reduction of associations with clinical outcomes, including VTE. Fourth, although Mendelian randomization can strengthen causal inference, limitations such as horizontal pleiotropy, linkage disequilibrium, and heterogeneity of instrument effects cannot be completely ruled out. Further functional validation is needed to confirm the biological relevance of the identified proteins. Finally, the ProteinRS was developed and evaluated within a single cohort. While internal cross-validation was performed, external validation in independent and more diverse populations is essential to assess its generalizability and clinical applicability.

## Conclusions

In conclusion, this study integrates genetics and proteomics to uncover key proteins and pathways involved in VTE. We identified proteins with both strong associations and potential causal roles and demonstrated that a protein-based risk score can improve prediction beyond clinical factors. These findings highlight the value of proteomics in understanding VTE biology and improving risk stratification, with potential applications in precision prevention and future therapeutic development.

## Methods

### Study Cohort

This study included 44,138 individuals from the UK Biobank with available proteomics measurements and genotype data. Individuals with discrepancies between self-reported and genetically determined sex were not classified among the cases. Participants where history of VTE or PE was only based on self-reports, were not considered as cases to ensure accurate classification. Since the PRS_VTE_ was calculated based on a GWAS conducted on individuals of European ancestry, only participants of European descent were included to ensure compatibility and reduce population stratification. Additionally, to focus on incident cases of VTE risk, participants with a prior diagnosis of VTE or PE at the time of enrollment were excluded from the analysis.

### Polygenic Risk Score (PRS)

PRS weights were obtained from a recent large-scale genome-wide association study (GWAS) by Ghouse et al.^16^, available through the PGS Catalog (PGS ID: PGS003332)^70^. This PRS was constructed using data from 57,467 VTE cases and 1,006,954 controls, with UK Biobank (UKB) samples intentionally excluded during the calculation to avoid potential biases. PRS-CS, a Bayesian-based method, was employed to model linkage disequilibrium (LD) using an external reference set (1000 Genomes European ancestry) and a continuous shrinkage prior to SNP effect sizes. The weights were applied to the UK Biobank cohort, and a PRS was calculated for each individual using PLINK^71^. The characteristics of the PRS, and its ability to predict VTE, have previously been described in detail^16^.

### UKB-PPP Plasma Proteomics Data

The plasma proteomics data for this study was obtained from the UK Biobank Pharma Proteomics Project (UKB-PPP), a collaboration between the UK Biobank and several biopharmaceutical companies, providing proteomic profiles for 54,219 participants. Using the antibody-based Olink Explore 3072 platform, this dataset quantifies levels of 2,923 proteins across diverse biological pathways, with each protein’s expression level reported in normalized units on a log2 scale^20,72,73^. Proteins with missing measurements in more than 20% of participants were excluded from subsequent analyses, resulting in a final set of 2,911 eligible proteins.

### Pathway Enrichment

To elucidate the biological implications of proteins associated with PRS_VTE_, we performed Gene Ontology (GO)^74^ enrichment analysis using the gost() function from the gprofiler2 package^75^. As input, we used the unranked list of proteins significantly associated with PRS_VTE_ (FDR-adjusted *P* value < 0.01). We specified the organism as human (“hsapiens”) and focused on the three GO categories: Biological Process (GO: BP), Molecular Function (GO: MF), and Cellular Component (GO: CC). We customized the background to only include the measured proteins in the present study. Statistical significance was determined using FDR correction, and all other parameters were set as default. Tissue-expression enrichment was performed with Enrichr^76^ (ARCHS4 Tissues RNA-seq library^77^, default settings). Significance was assessed by Fisher’s exact test with FDR correction.

### Mendelian Randomization

Mendelian Randomization is a method that uses genetic variation to identify potential causal relationships between exposures and outcomes^78^. We conducted two-sample MR analysis to investigate the causal associations between 56 proteins and VTE. These fifty-six proteins were identified to be both significantly associated with the PRS_VTE_ and associated with time-to-VTE event in Cox proportional hazards regressions. The MR analysis is bidirectional:(1) assessing the casual effects of protein levels on VTE risks, and (2) assessing the VTE genetic liability influencing protein expression levels.

Genetic instruments for each protein were identified from genome-wide significant single-nucleotide polymorphisms in a UK Biobank based GWAS on plasma protein levels^73^.

Specifically, cis-acting protein quantitative trait loci (cis-pQTLs) were selected as instrumental variables (IVs). SNPs were defined as cis-pQTLs if located within ± 1 megabase (Mb) of the transcription start site of the corresponding protein-coding gene. GWAS summary statistics were initially filtered at a genome-wide significance threshold (P < 5×10^−8^). If no cis-pQTLs met this threshold, we relaxed the threshold to P < 1×10^−6^ to retain informative SNPs. Instruments were then pruned to minimize linkage disequilibrium (LD), applying an R² threshold of 0.1 and a clumping window of 10 Mb. SNPs lacking genome coordinates, failing LD clumping, or having week instrument strength (mean F-statistic < 10) were excluded from the analysis. The outcome data for VTE were derived from the GWAS summary statistics published by Ghouse et al^16^. The VTE GWAS summary statistic were originally reported on GRCh37/hg19. We converted all variant coordinates to GRCh38/hg38 with the UVSC LiftOver tool (hg19-to-hg38 chain)^79^, retaining only variants that mapped uniquely. Outcome and exposure dataset were harmonized to ensure allele consistency. We used the inverse variance weighted (IVW) method as the primary analysis, providing causal effect estimates. For proteins with only one valid SNP after harmonization, we used the Wald ratio method. Sensitivity analyses were performed using the intercept of an MR-Egger regression to assess potential directional pleiotropy, and Cochran’s Q statistic to evaluate heterogeneity among the genetic instruments. These sensitivity analyses were reported only for proteins with sufficient number of valid SNPs remaining (≥2 SNPs for heterogeneity test, ≥3 SNPs for pleiotropy test). A log file of inclusion status, filtering thresholds, and reasons for protein exclusion during MR analysis is provided in **Supplementary Table 6**.

The analyses were conducted using R packages TwoSampleMR^80^ and MendelianRandomization^81^. We adjusted for multiple testing using an FDR correction.

### Protein-based Risk Score (ProteinRS) Construction

We constructed three LASSO regression models to predict incident VTE: (1) Protein-only model using the proteins selected by LASSO; (2) Clinical model including the selected clinical variables with non-zero coefficients (sex, smoking status, age at inclusion, BMI, diastolic blood pressure, systolic blood pressure, estimated glomerular filtration rate (eGFR), and triglycerides); (3) Combined model integrating both selected clinical risk factors and proteins. We then developed a protein-based risk score (ProteinRS) using the selected proteins from the protein-only model to predict the risk of incident VTE by plasma proteomics data.

The study cohort was randomly divided into a training dataset (70%, n = 30,897) and a testing dataset (30%, n = 13,241). Within the training set, LASSO regression models by the glmnet package^82^ in R were fitted for protein-only data, clinical-only data, and a combined dataset integrating both proteins and clinical variables. Missing protein values were imputed using k-nearest neighbor (k=10) imputation^83^, with imputation restricted to proteins with < 20% missingness. After imputation, all protein values were standardized to ensure comparability between proteins with differing expression ranges. Clinical variables considered for inclusion were age, sex, BMI, smoking status, cholesterol, LDL, HDL, triglycerides, eGFR, systolic blood pressure, and diastolic blood pressure. Those with missing data (e.g., smoking status, cholesterol levels, LDL, HDL, triglycerides, eGFR, and blood pressure measures) were imputed using linear regression models adjusted for sex, age at inclusion, BMI, batch effects, and the first ten genetic principal components. Incident VTE (coded as 1 if participants experienced either incident DVT or incident PE or both) was used as the outcome. Each model utilized 10-fold cross-validation, optimizing parameters based on maximizing the AUC. Optimal regularization parameters (λ) were determined by the one-standard-error rule (λ se) for the protein-only and combined models, and by minimum lambda (λmin) for the clinical-only model. We then compared three LASSO regression models refit using the optimal λ. Model performance was evaluated using the AUC, and pairwise comparisons between models were conducted using DeLong’s test^84^.

The selected protein features from the optimal protein-only LASSO model were subsequently extracted, retaining those with non-zero coefficients as predictors. These coefficients represented the relative importance of each protein biomarker and were used as weights when constructing the ProteinRS. We calculated the ProteinRS for each individual in the testing set by multiplying the expression level of each selected protein by its corresponding LASSO regression coefficient and summing these values across all proteins. The distribution of the ProteinRS among incident VTE cases and controls was visualized using density plots to illustrate differences between these groups.

To further evaluate the predictive ability of the ProteinRS, we stratified the study cohort into three risk groups based on ProteinRS percentiles: below the 20^th^ percentile, 20-80^th^ percentile (reference), and above the 80^th^ percentile. Cumulative incidence curves for VTE events within these groups were generated using the prodlim package^85^ in R, accounting for competing risks such as death and censoring. Cox proportional hazards models adjusted for clinical covariates (age, sex, BMI, smoking status, cholesterol, LDL, HDL, triglycerides, systolic blood pressure, and diastolic blood pressure) were used to estimate HRs and confidence intervals for incident VTE across groups. We then evaluated the risk stratification ability of extreme ProteinRS by defining binary indicators for individuals in the top 10% and top 1% of the ProteinRS distribution. Separate Cox models were fitted for each group to estimate HRs for VTE, adjusting for batch, age, sex, BMI, eGFR, and PC1-10. Results were visualized in a forest plot using the Publish package with log-scale HR. Additionally, we calculated incidence rates per 1,000 person-years within deciles of the ProteinRS. Incidence rates were visualized on a logarithmic scale to examine the relationship between ProteinRS percentile and VTE.

### Statistical Analyses

The two-step statistical analysis served as an observational mediation analysis prior to MR analysis. All statistical analyses were conducted using R version 4.2.0^86^. To address multiple testing, Click or tap here to enter text.False Discovery Rate (FDR) corrections^88^ adjustment was applied depending on the analysis.

### PRS_VTE_ and Protein Association

To assess the association between the PRS_VTE_ and individual protein expression levels, we performed a series of multivariable linear regressions, treating each protein as the response variable. The regression model included the PRS_VTE_ as the primary predictor, with additional covariates (Batch, sex, age incl, BMI, eGFR, and the top 10 genotype principal components— PC1 to PC10). FDR correction was applied to adjust for multiple comparisons, and an adjusted *P* value < 0.01 was considered to be significant.

### Protein Time-to-Event Association

To evaluate the association between protein levels and the hazard of VTE over time, Cox regression analyses were conducted using the survival package in R^89^ on all 2911 measured proteins. The index date was set to be the time of enrollment in the UK Biobank Participants were followed from the date of enrollment in the UK Biobank (index date) to the first VTE event, death, or end of follow-up (censor), whichever came first. Each protein was treated as a predictor variable, with the time to VTE as the response variable. Models were adjusted for plasma proteomics batch, sex, age incl, BMI, eGFR, and the top 10 genotype principal components to control for confounding. Hazard ratios (HRs) with 95% confidence intervals and *P* values were extracted. FDR adjustment was applied to correct for multiple comparisons, and statistically significant results were identified at an adjusted *P* value threshold of 0.01.

To visualize the relationship between the protein hazard ratios obtained from Cox regression and the effect estimates derived from linear regression analyses assessing protein associations with PRS, we constructed a scatter plot. Proteins identified as significant in either analysis were highlighted. Specifically, proteins significantly associated with time-to-event (Cox regression FDR-corrected *P* value < 0.01) and/or significantly associated with PRS_VTE_ (linear regression FDR-corrected *P* value < 0.01) were indicated with distinct colors. The x-axis displayed hazard ratios (HR) from the Cox regression on a logarithmic scale, reflecting risk per protein expression standard deviation increase. The y-axis is the linear regression estimates, representing changes in protein expression per PRS_VTE_ standard deviation increase. Reference lines were drawn at HR = 1 (vertical) and estimate = 0 (horizontal) for clarity.

## Spline Analysis

To further evaluate potential non-linear associations, and illustrate HR across protein expression ranges, we constructed penalized spline representations (psplines) of the Cox regression models. Each protein was analyzed using a Cox proportional hazards model with splines to explore non-linear effects. The formula included the protein expression levels represented by penalized splines with 4 degrees of freedom and the same covariates as in the Cox model. The *P* values for non-linearity were adjusted by FDR correction, with FDR < 0.01 considered to be significant. Plots of the spline functions were generated to visualize the relationship between protein expression and the HRs, including 95% confidence intervals.

## Data Availability

Access to the UK Biobank is available for bona-fide researchers through https://www.ukbiobank.ac.uk/enable-your-research/apply-for-access

## Extended data figures

**Extended Data Fig. 1:**
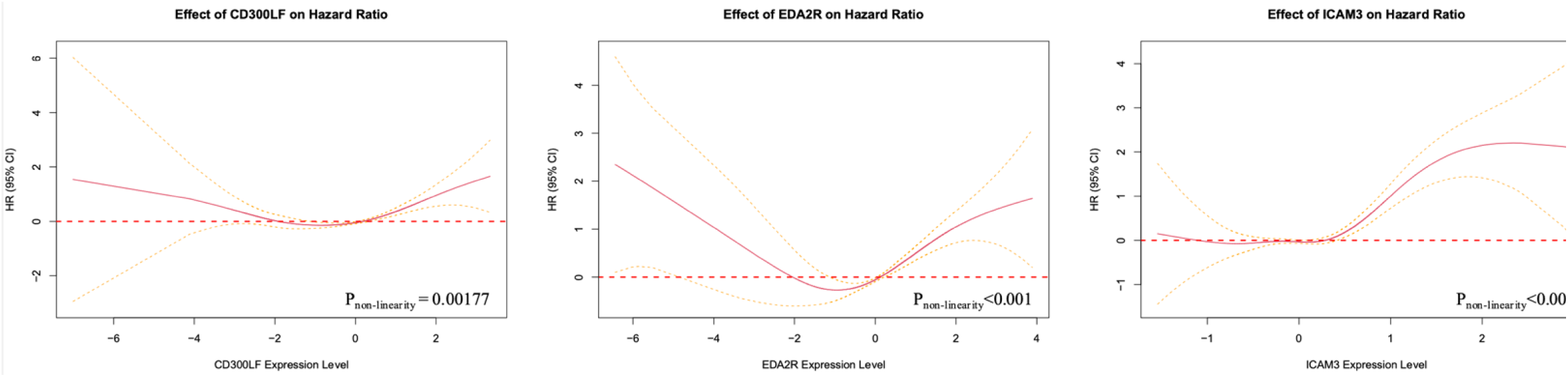
Non-linear effects of protein expression on HRs. Penalized smoothing splines showing the relationship between protein expression levels and HRs for four proteins with significant non-linear effects (FDR < 0.001): CD300LF, EDA2R, and ICAM3. The solid red lines represent estimated HRs, and the dashed yellow lines indicate 95% confidence intervals.

**Extended Data Fig. 2.**
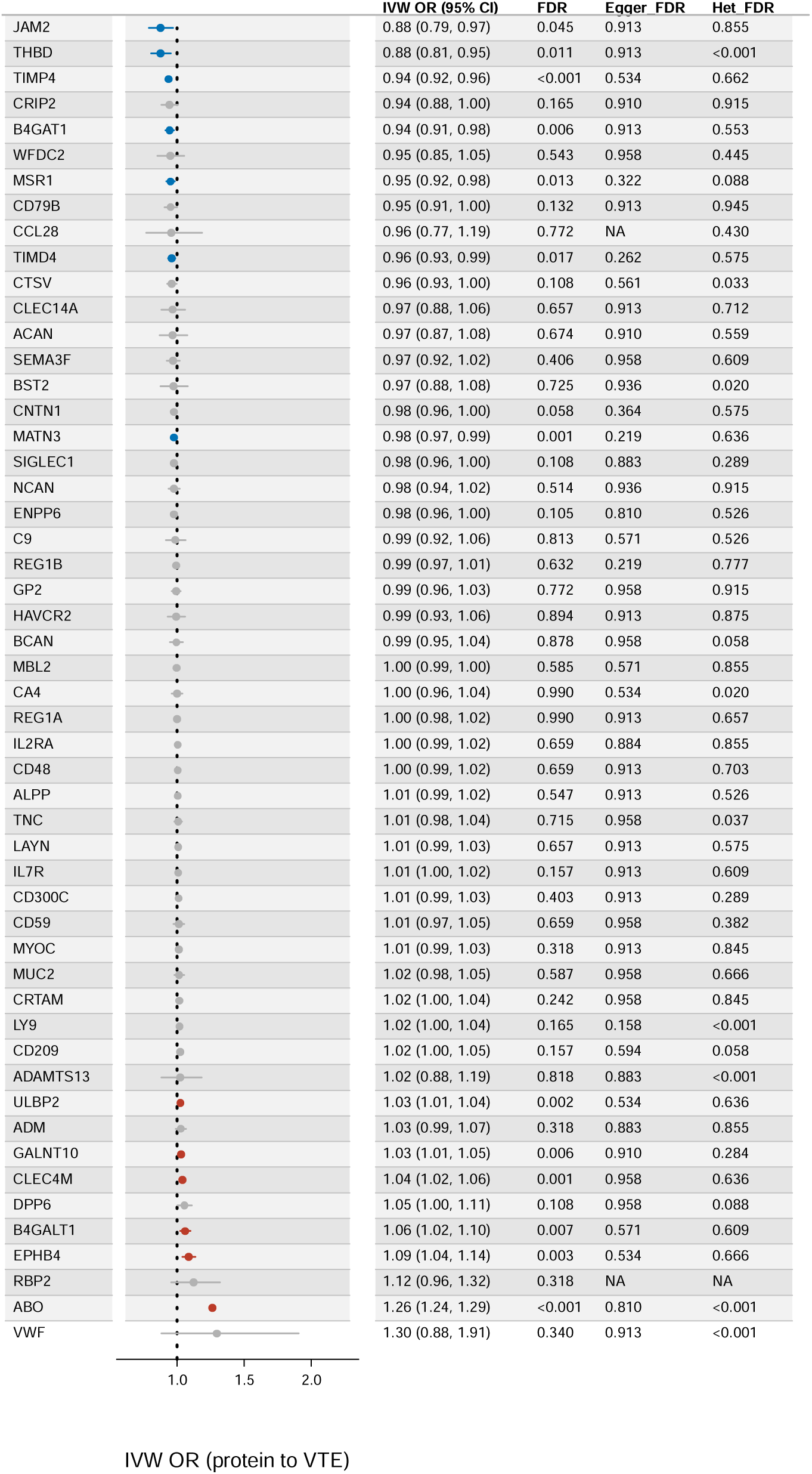
Comprehensive Mendelian randomization results for proteins on VTE. Odds ratios (ORs) and 95% confidence intervals (CIs) from inverse-variance weighted (IVW) MR analyses represent causal estimates of protein levels on VTE risk. Blue indicates a higher protein level associated with lower VTE risk, red indicates a higher protein level associated with higher VTE risk, and grey represents proteins without significant casual relationships. Cochran’s Q statistic (heterogeneity) and MR-Egger intercept (pleiotropy) test results are provided as False Discovery Rate (FDR)-adjusted *P* values (FDR).

**Extended Data Fig. 3.**
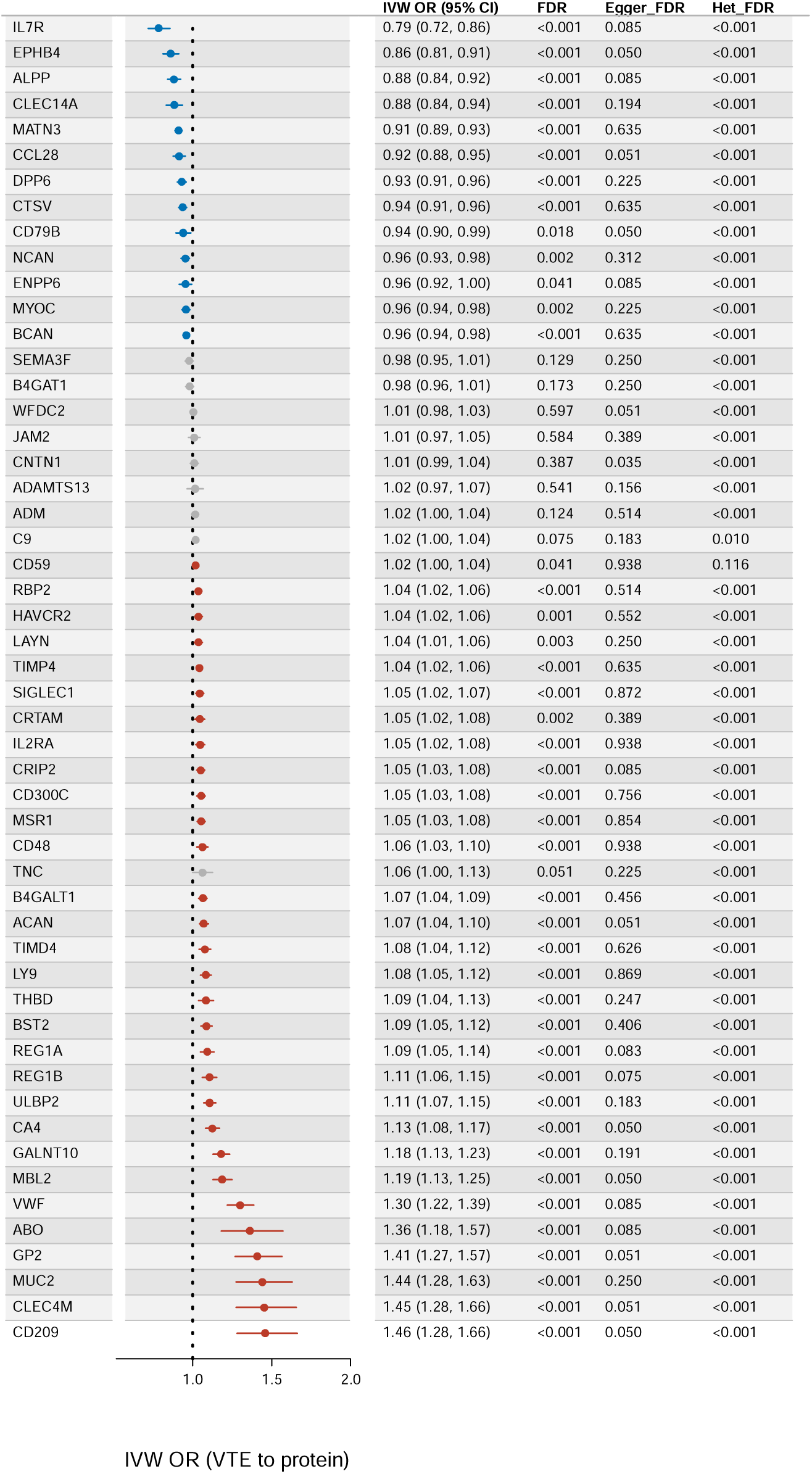
Comprehensive Mendelian randomization results for VTE risk on proteins. Odds ratios (ORs) and 95% confidence intervals (CIs) from inverse-variance weighted (IVW) MR analyses represent causal estimates of VTE genetic predisposition on protein expression levels. Blue dots indicate proteins with significantly lower levels associated with increased VTE genetic liability, red dots indicate proteins with significantly higher levels associated with increased VTE genetic liability, and grey represents proteins without significant casual relationships.

